# Arachnoid Granulation Morphologies Associate with β-Amyloid and Tau Pathology in Older Adults

**DOI:** 10.1101/2025.09.30.25336940

**Authors:** Rashi I. Mehta, Tianhao Wang, Aubrey Lewis, Claire E. Walker, Caroline Gebczak, Lisa L. Barnes, David A. Bennett, Rupal I. Mehta

## Abstract

**INTRODUCTION:** Arachnoid granulations (AG) enlarge with age, yet limited studies evaluate AG in neurodegeneration. Here, we investigate associations of AG morphology with Alzheimer’s disease (AD) pathology.

**METHODS:** Macroscopic AG properties were systematically evaluated along dorsal postmortem brain specimens from older adults (n=882). Regression models were used to analyze their associations with AD neuropathologic indices, controlling for demographic factors.

**RESULTS:** Participants died at mean age of 90.62 (SD=6.98) years. β-amyloid (Odds ratio/OR, 0.79 [95% CI, 0.66-0.95]) and neurofibrillary tangles (OR, 0.84 [95% CI, 0.71-0.98]) inversely associated with global AG count. Similarly, β-amyloid (OR, 0.78 [95% CI, 0.65-0.92]) and B (Braak) score (OR, 0.80 [95% CI, 0.66-0.96]) inversely associated with global AG patch count.

**DISCUSSION:** β-amyloid and tau pathology are associated with AG morphologies. Future studies should explore the mechanisms underlying these associations across disease stage and demographic factors.

## Introduction

Arachnoid granulations (AG) are localized exophytic protrusions of the brain’s middle meningeal layer, i.e., the arachnoid mater, that function in cerebrospinal fluid (CSF) resorption [Brunori et al., 1993; Pollay et al., 2010]. These structures were described as early as the sixteenth century [Brunori et al., 1993; Pollay et al., 2010], yet few reports systematically investigate AG in humans. Moreover, varied descriptions of AG morphology in lower animals, including sheep and monkeys, have led to conflicting concepts regarding their structure and function in humans [Gomez et al., 1973; Gomez et al., 1974]. Thus, the study of AG is a critical gap area in clinical anatomy and physiology.

Despite known roles of AG in intracranial fluid clearance, the associations of AG with diseases are largely unprobed [Welch and Friedman, 1960; Wolpow and Schaumburg, 1972]. This has been due, in large part, to confusion regarding their typical morphologies. Recent work by our group demonstrated marked heterogeneity in AG anatomy across aging [Shah et al., 2023], suggesting potential associations of AG with age-related pathologies. However, this prior analysis was limited to postmortem investigation of AG along frontal cerebral cortical regions of a limited number of hospital decedents devoid of intracranial pathology. Thus, additional comprehensive clinicopathologic analyses of AG, including characterization of their morphology across brain regions and their associations with diseases are needed.

Alzheimer’s disease (AD), the most prevalent form of dementia and a major global health challenge [Alzheimer’s Association, 2025], is characterized by the intracerebral aggregation of β-amyloid and paired helical filament-tau (PHF-Tau) proteins, i.e., biomarkers recognized in the “A/T/N” classification [Jack Jr. et al., 2016]. Though its cause remains uncertain, β-amyloid and PHF-Tau (i.e., A/T) accumulation, possibly a result of compromised clearance mechanisms, is known to lead to neurodegeneration [Jack Jr. et al., 2018; Mehta and Schneider, 2021]. Given the predominance of AG along parasagittal brain borders, presumed sites of fluid and molecular egress, and the importance of intracranial fluid physiology in brain maintenance and diseases [Louveau et al., 2021; Da Mesquita et al., 2018], we theorize that AG serve critical roles in cerebral protein clearance. To investigate the potential impact of AG on AD pathology, we systematically studied the macroscopic properties of AG along dorsal brain surfaces of individuals who were followed in longitudinal studies of aging and found that various dorsal AG morphologic properties associate with postmortem intracerebral A/T indices.

## Materials and Methods

### Participants and Brain Specimens

Brain specimens derived from one of five ongoing community-based longitudinal, clinical-pathologic studies of aging at the Rush Alzheimer’s Disease Center: The Religious Orders Study (ROS) [Bennett et al., 2006; Bennett et al., 2018], the Rush Memory and Aging Project (MAP) [Bennett et al., 2018], the Minority Aging Research Study (MARS) [Barnes et al., 2012], the African American Clinical Core (AACore) [Schneider et al., 2009], and the Latino Core (LATC) [Marquez et al., 2020]. ROS began enrolling Catholic clergy (nuns, priests, and brothers) in 1994 from over 40 monasteries or convents across the United States with only a minority of autopsies at the RADC, whereas MAP began enrolling older laypersons from the Chicago region in 1997, MARS began recruiting minoritized seniors from a variety of community-based settings in the metropolitan Chicago area and outlying suburbs beginning in 2004, and AACore and LATC were initiated subsequently [Barnes et al., 2012; Bennett et al., 2018; Marquez et al., 2020; Schneider et al., 2009]. All studies were approved by an Institutional Review Board of the Rush University Medical Center. All participants sign informed and repository consents and all ROS and MAP and many from the other cohorts sign an Anatomical Gift Act for brain donation. Included in the studies were adults with no known dementia at baseline who consented to undergo annual clinical evaluations. Across all five cohorts, the autopsy rate is approximately 85%, and additional details regarding study designs and components have been published previously [Bennett et al., 2018]. The studies have harmonized methods for enrollment and procedures including data collection and summarization, allowing for combined datasets to be analyzed [Barnes et al., 2012; Marquez et al., 2020; Schneider et al., 2009].

### Brain Autopsy and Tissue Processing

Brain autopsies were performed by trained staff who were blinded to demographic, clinical, and other neuropathologic information. As previously described, a standardized procedure was used for autopsy procedures following variable postmortem intervals (PMI) [Bennett et al, 2018]. In brief, the brains of deceased participants were systematically and carefully extracted, weighed, and photographed in various precise planes after intracranial removal and brain images were stored in an internal database. For processing, one cerebral hemisphere was frozen while the other cerebral hemisphere was sliced into 1 cm coronal slabs that were fixed in 4% paraformaldehyde for 3 to 21 days. Subsequently, the fixed brain slabs were examined to evaluate macroscopic tissue lesions and diagnostic tissue blocks were dissected from standardized brain regions (i.e., at least nine brain regions in total, encompassing middle frontal, middle temporal, amygdala, hippocampus, inferior parietal, occipital, basal ganglia, thalamus, and midbrain) as well as areas with grossly visible brain lesions. Collected tissue blocks were paraffin embedded and serially sliced into 6 μm sections that were mounted on glass slides. Hematoxylin and eosin (H&E)-stained sections were prepared from each block and modified Bielschowsky silver stain and β-amyloid immunohistochemistry slide preparations using anti-β-amyloid antibody (4G8, 1:9000, BioLegend #800711, San Diego, CA, USA); 6F/3D, 1:50, Dako #M0872, Carpinteria, CA, USA; 10D5, 1:600, Elan Pharmaceuticals, San Francisco, CA, USA) were prepared from designated blocks, according to consensus protocols [Hyman et al., 2012]. Diagnostic examination was completed in a systematic manner and was overseen by board-certified neuropathologists who were blinded to all demographic, genetic, and clinical data.

### Evaluation of Macroscopic AG Properties

Bilateral frontal, parietal, occipital, and cerebellar regions were examined for the presence or absence of AG. Data on AG characteristics were acquired from a given brain region only if a dorsal brain photograph was available in the database and if the leptomeninges were present and intact on the brain surface. To ensure systematic assessment of AG, brains with obscuration by hemorrhage, bone dust, bonesaw damage and/or other lesions that obstructed visualization of the dorsal brain surface were excluded. Data collection on AG properties was overseen by a licensed pathologist who was experienced and board-certified in both anatomic pathology and neuropathology. Each brain photograph was reviewed by at least two reviewers who underwent extensive training with the neuropathologist. For each brain specimen, the presence, number (i.e., count), size, location(s), and morphologic type(s) of AG were assessed on bilateral brain hemispheres. AG properties were individually evaluated for each cerebral lobe (i.e., frontal, parietal, and occipital) and the cerebellar hemisphere. For AG count, AG were semiquantitatively scored using an ordinal scale: 0 (none), 1 (1 to 5), 2 (6 to 10), 3 (11 to 15), 4 (16 to 20), 5 (21 to 50), 6 (51 to 100), 7 (101 to 200), or 8 (over 200) per brain region. The maximum dimension (in mm) and location of the ten largest AG per brain specimen were recorded. For AG type, AG were characterized as solitary, linear, or patch morphology. The solitary type was characterized by singular or individually scattered AG, whereas linear type included conglomerated AG that were present in elongated strands (at least 1 cm long and less than 5 mm wide) that were parallel to the midsagittal plane. Patch type included conglomerated AG (5 or more contiguous structures) in which the area occupied at least a 5 mm by 5 mm area. For AG count, the mean semiquantitative score between the left and right sides was used for analyses. For maximum AG size, the largest AG dimension across bilateral lobes or hemispheres was used. For AG type, each AG morphology (i.e., solitary, linear, or patch type) was summarized as present or absent and the number of patch type AG (i.e., “patch count”) was recorded for each brain region using an ordinal score representing 0 (none), 1 (singular patch), or 2 (more than 1 patch). For analyses of patch count, the mean of left and right lobes (or hemispheres) were used. Note that for this study, data were not analyzed with regard to laterality.

### Evaluation of AD Pathology and A/T Indices

The presence of diffuse plaques, neuritic plaques, and neurofibrillary tangles was evaluated on modified Bielschowsky silver stain. Each lesion type was also counted in the entorhinal, hippocampal CA1, middle frontal, superior temporal, and angular gyrus regions exhibiting the highest count in a 1 mm^2^ area. Per National Institute on Aging–Alzheimer Association criteria [Hyman et al., 2012], Alzheimer’s disease neuropathologic change (ADNC) was used to render a pathologic diagnosis of AD and disease severity was graded from 0 (no ADNC) to 3 (high ADNC) [Montine et al., 2012]. For the study, an ordinal variable indicating none, low, intermediate, or high ADNC was used along with a quantitative diffuse plaque variable that reflected the intracerebral diffuse plaque count. Quantitative neuritic plaque, neurofibrillary tangle, and combined neuritic plaque and neurofibrillary tangle variables (based on silver stain and reflecting the count of each individual lesion) and other neuropathologic A/T indices (based on immunohistochemical, i.e., “IHC” staining and reflecting the intracerebral burden of IHC label) were also used and were square root transformed due to positive skewness. Total plaques (a combined measure of diffuse plaques and neuritic plaques), ADNC score, A score (a measure of Thal level, i.e., topographical intraparenchymal β-amyloid score), B score (a measure of Braak level, i.e., topographical neurofibrillary tangle score), and C score (a measure of neocortical neuritic plaque density) were additionally analyzed [Montine et al., 2012].

### APOE Genotypes

For determination of *APOE* genotypes, DNA was extracted from peripheral blood mononuclear cells or brain, and genotyping was performed utilizing high-throughput sequencing of codon 112 (position 3937) and codon 158 (position 4075) of exon 4 of the *APOE* gene on chromosome 19 [Yu et al., 2017]. For analyses, the *APOE* genotypes were grouped by presence or absence of an *APOE* ε4 allele.

### Statistical Analysis

For analyses, dichotomous variables representing the presence or absence of solitary, linear, or patch type AG were used. For AG size, a continuous measure was used, whereas ordinal scores were used for AG count and patch count. Chi-square and t tests were used to compare demographic and AG properties. Multiple logistic regression analyses were used to evaluate the associations of AG properties with the following: (1) ADNC score [Montine et al., 2012]; (2) individual neuropathologic lesions [diffuse plaques, neuritic plaques, neurofibrillary tangles, β-amyloid, and PHF-Tau]; and (3) combined neuropathologic lesions or summary scores [total plaques and A/B/C scores]. Logistic regression models were used to test the associations of AG type with A/T neuropathologic indices. Ordinal regression models were used to test the associations of AG count and AG patch count with A/T neuropathologic indices. Linear regression models were used to test the associations of AG size with A/T neuropathologic indices. Using these models, we also checked the potential interaction effects of AG properties with each other and with race (i.e., non-Latino Blacks versus non-Latino Whites). In secondary analyses, we adjusted the primary models for brain weight, lobe length, PMI, and presence of *APOE* □4 allele. All models controlled for age at death, sex, and education. Analyses were carried out using SAS version 9.4 (SAS Institute). A threshold of p<0.05 was used for statistical significance throughout.

## Data availability

Information regarding the data used and the parent studies description can be accessed on the Rush Alzheimer’s Disease Center Research Resource Sharing Hub at https://www.radc.rush.edu. Following review of a submitted data request, approved applicants with a Data Use Agreement can receive data.

## Results

Of enrolled participants, 649 MAP, 134 ROS, 70 MARS, 27 AA, and 2 LATC participants died, underwent autopsy, and had completed autopsy reports with unobstructed dorsal brain photographs available at the time of this analysis. Thus, 882 brains from deceased community-dwelling older persons were studied. The eligible participants included 326 participants without a pathologic diagnosis of AD (i.e., with no or low ADNC) and 556 participants with a pathologic diagnosis of AD (i.e., intermediate or high ADNC). *APOE* genotype was available in 808 (91.61%) of these participants. Demographic and *APOE* characteristics are summarized in the entire group and by AD status in Table 1 (see Tables S1 and S2 for summaries by race). Briefly, participants died on average at the age of 90.6 ± 7.0 years, were mostly women (n=660, 75%) and highly educated, and 208 (23.6%) had an *APOE* ε4 allele. The relationships of AG properties with each other and with demographic factors and neuropathologic changes are summarized below.

**Table 1.**
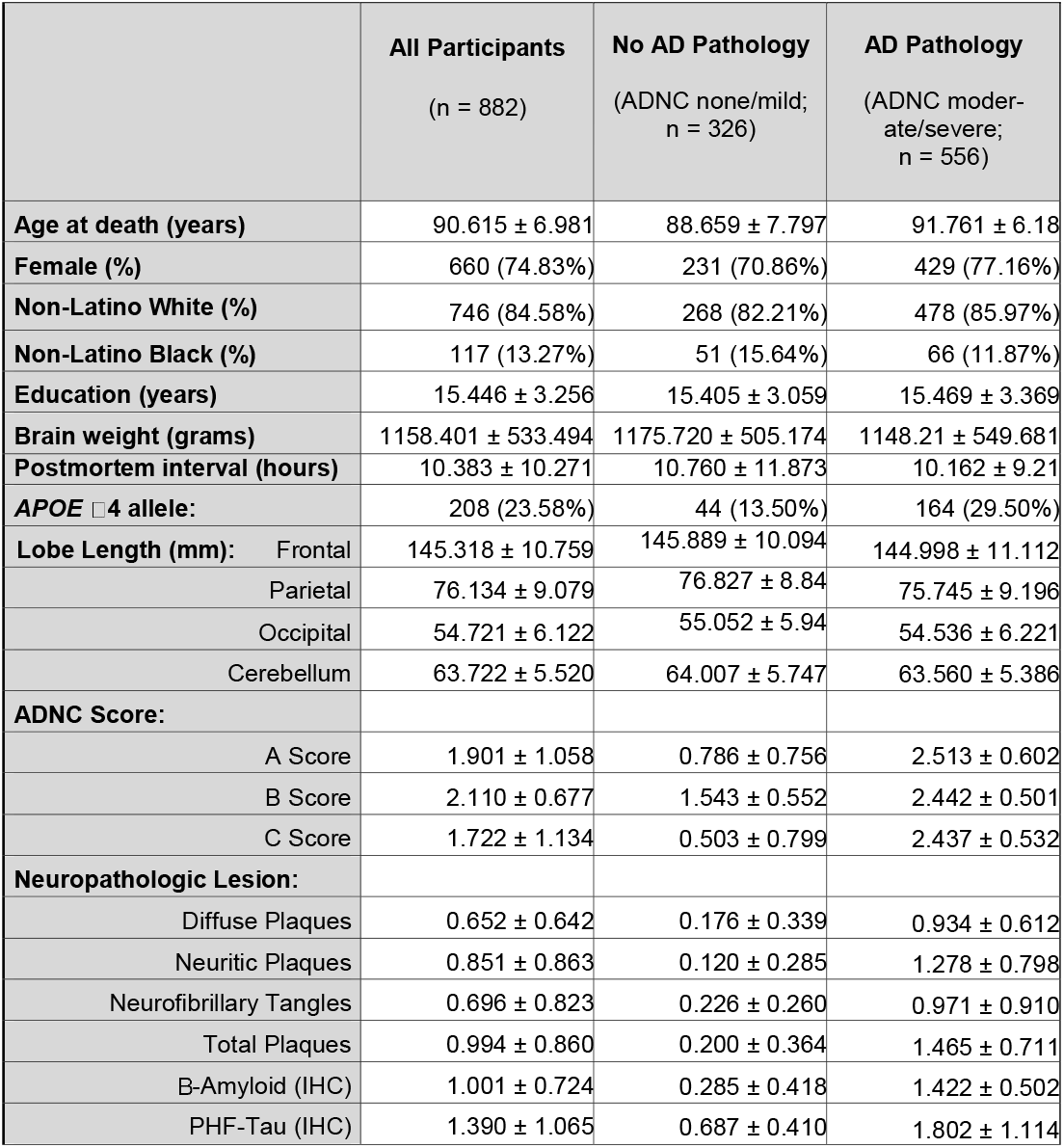
Demographic and neuropathologic characteristics of participants and postmortem brain specimens (n = 882).

|  | All Participants<br>(n = 882) | No AD Pathology<br>(ADNC none/mild;<br>n = 326) | AD Pathology<br>(ADNC moderate/severe;<br>n = 556) |
| --- | --- | --- | --- |
| Age at death (years) | 90.615 ± 6.981 | 88.659 ± 7.797 | 91.761 ± 6.18 |
| Female (%) | 660 (74.83%) | 231 (70.86%) | 429 (77.16%) |
| Non-Latino White (%) | 746 (84.58%) | 268 (82.21%) | 478 (85.97%) |
| Non-Latino Black (%) | 117 (13.27%) | 51 (15.64%) | 66 (11.87%) |
| Education (years) | 15.446 ± 3.256 | 15.405 ± 3.059 | 15.469 ± 3.369 |
| Brain weight (grams) | 1158.401 ± 533.494 | 1175.720 ± 505.174 | 1148.21 ± 549.681 |
| Postmortem interval (hours) | 10.383 ± 10.271 | 10.760 ± 11.873 | 10.162 ± 9.21 |
| APOE ε4 allele: | 208 (23.58%) | 44 (13.50%) | 164 (29.50%) |
| Lobe Length (mm): Frontal | 145.318 ± 10.759 | 145.889 ± 10.094 | 144.998 ± 11.112 |
| Parietal | 76.134 ± 9.079 | 76.827 ± 8.84 | 75.745 ± 9.196 |
| Occipital | 54.721 ± 6.122 | 55.052 ± 5.94 | 54.536 ± 6.221 |
| Cerebellum | 63.722 ± 5.520 | 64.007 ± 5.747 | 63.560 ± 5.386 |
| ADNC Score: |  |  |  |
| A Score | 1.901 ± 1.058 | 0.786 ± 0.756 | 2.513 ± 0.602 |
| B Score | 2.110 ± 0.677 | 1.543 ± 0.552 | 2.442 ± 0.501 |
| C Score | 1.722 ± 1.134 | 0.503 ± 0.799 | 2.437 ± 0.532 |
| Neuropathologic Lesion: |  |  |  |
| Diffuse Plaques | 0.652 ± 0.642 | 0.176 ± 0.339 | 0.934 ± 0.612 |
| Neuritic Plaques | 0.851 ± 0.863 | 0.120 ± 0.285 | 1.278 ± 0.798 |
| Neurofibrillary Tangles | 0.696 ± 0.823 | 0.226 ± 0.260 | 0.971 ± 0.910 |
| Total Plaques | 0.994 ± 0.860 | 0.200 ± 0.364 | 1.465 ± 0.711 |
| B-Amyloid (IHC) | 1.001 ± 0.724 | 0.285 ± 0.418 | 1.422 ± 0.502 |
| PHF-Tau (IHC) | 1.390 ± 1.065 | 0.687 ± 0.410 | 1.802 ± 1.114 |

### AG Properties

Nearly all participants (880, 99.8%) exhibited presence of AG on the brain surface. The AG were present in variable number, size, distribution, and morphologic type at the dorsal brain surface. The overall presence and frequency of AG were highest in the frontal region and diminished gradually along the posterior brain convexity, being rare over the cerebellum (Fig 1A,B). Due to the low number of AG along the cerebellar hemispheres and their irregular obscuration by cerebral hemispheres, AG properties (apart from presence and count) were further studied in the frontal, parietal, and occipital regions and combined cerebrum (hereafter referred to as “brain”) regions, only. Representative images depicting AG morphologic types, including solitary, patch, and linear type are shown in Fig 1C. Overall, the solitary type was most frequent, followed by patch and then linear type (Table 2; Fig 1D). Overall, the AG size ranged from 1 mm to 5.7 cm (mean, 9.9 ± 4.2 mm). The frontal AG size correlated highly with parietal (corr=0.34, p=<0.0001) and occipital (corr=0.56, p=<0.0001) AG sizes. Overall, the brain AG size correlated highly with the brain AG number (corr=0.15, p=<0.001) (Fig 1E). Within lobar regions, the AG patch count correlated highly with the AG count (p=<0.0001 for all lobes), but not with AG size.

**Table 2.** AG characteristics in postmortem brain specimens (n = 882). *“Brain” represents composite dorsal cerebrum. **AG counts represent the mean of semiquantitative scores from the left and right sides.

|  | All Participants<br>(n = 882) | No AD Pathology<br>(ADNC none/mild;<br>n = 326) | AD Pathology<br>(ADNC moderate/severe;<br>n = 556) |
| --- | --- | --- | --- |
| **Brains with any AG (n, %)** |  |  |  |
| Frontal | 850 (96.37%) | 312 (95.71%) | 538 (96.76%) |
| Parietal | 840 (95.24%) | 308 (94.48%) | 532 (95.68%) |
| Occipital | 771 (87.41%) | 284 (87.12%) | 487 (87.59%) |
| Cerebellum | 303 (34.35%) | 105 (32.21%) | 198 (35.61%) |
| Brain* | 880 (99.77%) | 325 (99.69%) | 555 (99.82%) |
| **AG count \*\*** |  |  |  |
| Frontal | 3.926 ± 1.390 | 3.934 ± 1.422 | 3.921 ± 1.372 |
| Parietal | 3.157 ± 1.348 | 3.204 ± 1.406 | 3.13 ± 1.313 |
| Occipital | 0.93 ± 0.631 | 0.948 ± 0.638 | 0.92 ± 0.627 |
| Brain* | 2.073 ± 0.669 | 2.086 ± 0.671 | 2.065 ± 0.669 |
| **AG size (max, in mm)** |  |  |  |
| Frontal | 15.191 ± 8.111 | 14.967 ± 8.186 | 15.370 ± 8.059 |
| Parietal | 11.970 ± 6.837 | 11.746 ± 6.447 | 12.160 ± 7.160 |
| Occipital | 8.556 ± 5.975 | 9.192 ± 7.020 | 8.108 ± 5.174 |
| Brain* | 16.589 ± 8.028 | 16.374 ± 7.952 | 16.759 ± 8.096 |
| **Presence of diffuse type AG (n, %)** |  |  |  |
| Frontal | 849 (96.26%) | 312 (95.71%) | 537 (96.58%) |
| Parietal | 838 (95.01%) | 307 (94.17%) | 531 (95.50%) |
| Occipital | 771 (87.41%) | 284 (87.12%) | 487 (87.59%) |
| **Presence of patch type AG (n, %)** |  |  |  |
| Frontal | 489 (55.44%) | 187 (57.36%) | 302 (54.32%) |
| Parietal | 264 (29.93%) | 99 (30.37%) | 165 (29.68%) |
| Occipital | 31 (3.51%) | 12 (3.68%) | 19 (3.42%) |
| **Presence of linear type AG (n, %)** |  |  |  |
| Frontal | 169 (19.16%) | 59 (18.10%) | 110 (19.78%) |
| Parietal | 98 (11.11%) | 41 (12.58%) | 57 (10.25%) |
| Occipital | 3 (0.34%) | 1 (0.31%) | 2 (0.36%) |
| **AG patch Count (n)** |  |  |  |
| Frontal | 0.79 ± 0.77 | 0.80 ± 0.75 | 0.78 ± 0.78 |
| Parietal | 0.38 ± 0.61 | 0.41 ± 0.64 | 0.37 ± 0.59 |
| Occipital | 0.04 ± 0.20 | 0.04 ± 0.20 | 0.04 ± 0.20 |
| Brain* | 0.88 ± 0.77 | 0.90 ± 0.76 | 0.87 ± 0.77 |

**Figure 1.**
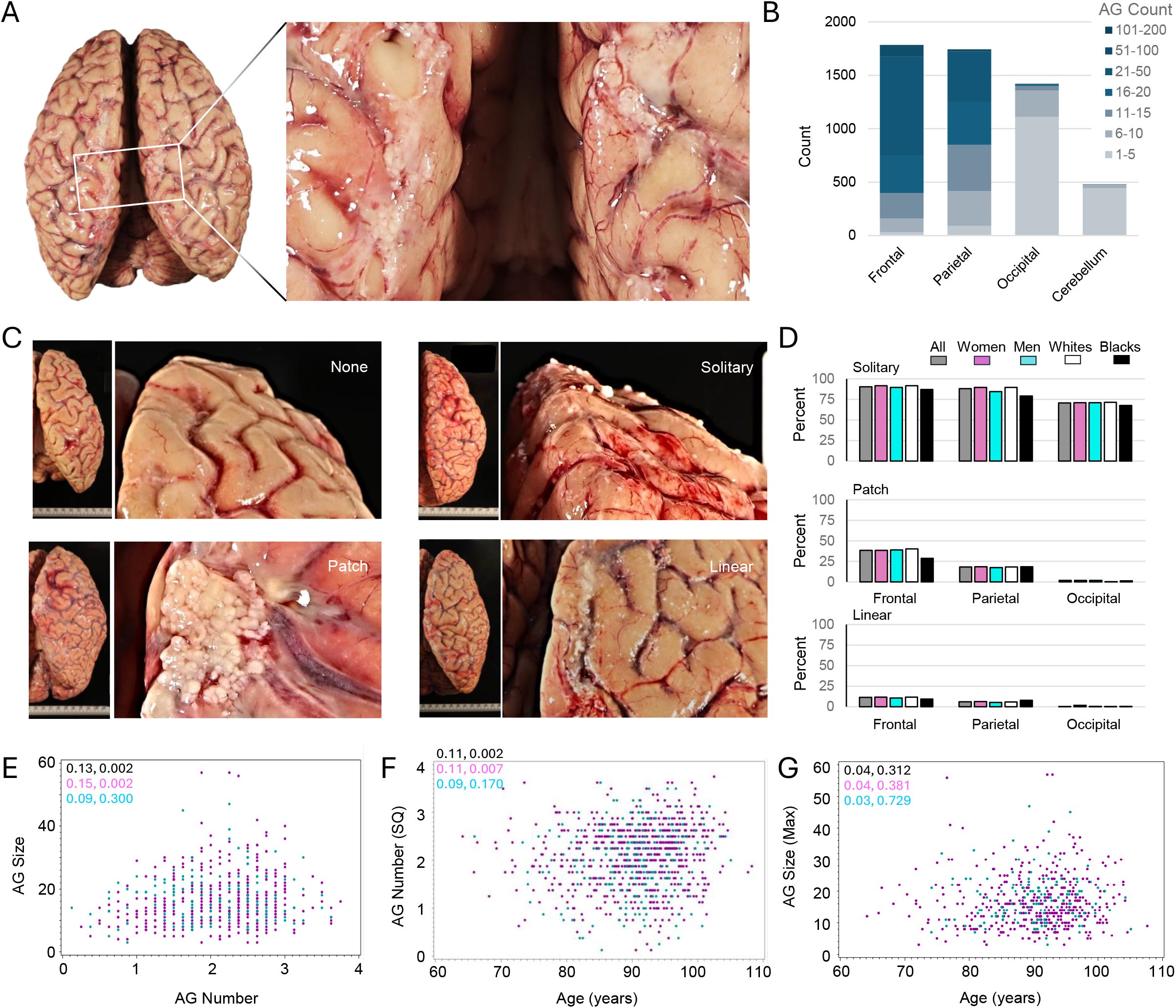
AG properties at the dorsal human brain surface. A banked postmortem photograph demonstrates counts, types, and distribution of AG at the dorsal brain surface **(A)**. As shown in the boxed area and enlarged inset, AG were prominent in the dorsal frontal and parietal regions. AG counts in different brain regions are summarized in **(B)**. Representative images depict different AG types **(C)** and the graphs summarize their numbers and locations in dorsal brain **(D)**. Scatterplots depict the relationship between AG count and AG size **(E)**, AG count and age at death **(F)**, and AG size and age at death **(G)**; correlation coefficients and p values are shown for the entire group (black) and subsets of men (cyan) and women (pink). n=882 brain specimens (in total).

### AG Properties, Age, and Brain Characteristics

The AG count in the frontal region (corr=0.09, p=0.006), parietal region (corr=0.09, p=0.007), cerebellar region (corr=0.07, p=0.034), and brain (corr=0.11, p=0.002) each correlated with age at death (Pearson correlation coefficient) (Fig 1F). However, AG count in the occipital region showed no association with age at death (corr=<0.01, p=0.992). Interestingly, AG size did not correlate with age at death in this cohort (Fig 1G), though the frontal lobe patch count (corr=0.09, p=0.007) and brain patch count (0.09, p=0.011) both correlated with age at death. AG count, AG size, and AG patch count did not correlate with brain weight or PMI in any brain region. In regression analyses adjusting for age at death, sex, and education, corresponding lobe length associated with AG count within the occipital lobe (OR, 1.03 [95% CI, 1.01–1.06]). The brain length associated with brain AG count (OR, 0.99 [95% CI, 0.98–1.00]) as well as presence of patch type AG in the composite brain region (OR, 0.99 [95% CI, 0.98–1.00]). Other AG properties were not associated with lobe lengths or brain length when adjusting for demographics.

### AG Count and A/T Indices

We next studied the associations of neuropathologic indices with AG count using regression models, controlling for demographics. We found that a higher brain β-amyloid load is associated with lower odds of high AG count in the frontal region (OR, 0.82 [95% CI, 0.69–0.98]), parietal region (OR, 0.75 [95% CI, 0.63–0.89]), occipital region (OR, 0.81 [95% CI, 0.67–0.97]), and brain (OR, 0.79 [95% CI, 0.66–0.95]). Similarly, a higher brain neurofibrillary tangle load is associated with lower odds of high AG count in the frontal region (OR, 0.85 [95% CI, 0.73– 1.00]), parietal region (OR, 0.84 [95% CI, 0.72–0.98]), occipital region (OR, 0.76 [95% CI, 0.65–0.90]), and brain (OR, 0.84 [95% CI, 0.71–0.98]). Multiple additional associations were found between neuropathologic indices and AG count. The odds of high AG count in the occipital region was lower among those with higher ADNC score (OR, 0.86 [95% CI, 0.75–0.99]), high neuritic plaque count (OR, 0.85 [95% CI, 0.73–0.99]), and high PHF-Tau burden (OR, 0.85 [95% CI, 0.75–0.97]). The odds of high AG count in the parietal region (OR, 0.77 [95% CI, 0.64– 0.93]) and occipital region (OR, 0.71 [95% CI, 0.58–0.88]) was lower among those with a high B score. The odds of high AG count in the parietal region was lower among those with high total plaque load (OR, 0.87 [95% CI, 0.75–1.00]), and the odds of high AG count in the brain was lower among those with high diffuse plaque load (OR, 0.80 [95% CI, 0.65–0.99]). The associations between AG count and various neuropathologic indices are summarized by brain region in Fig. 2.

**Figure 2.**
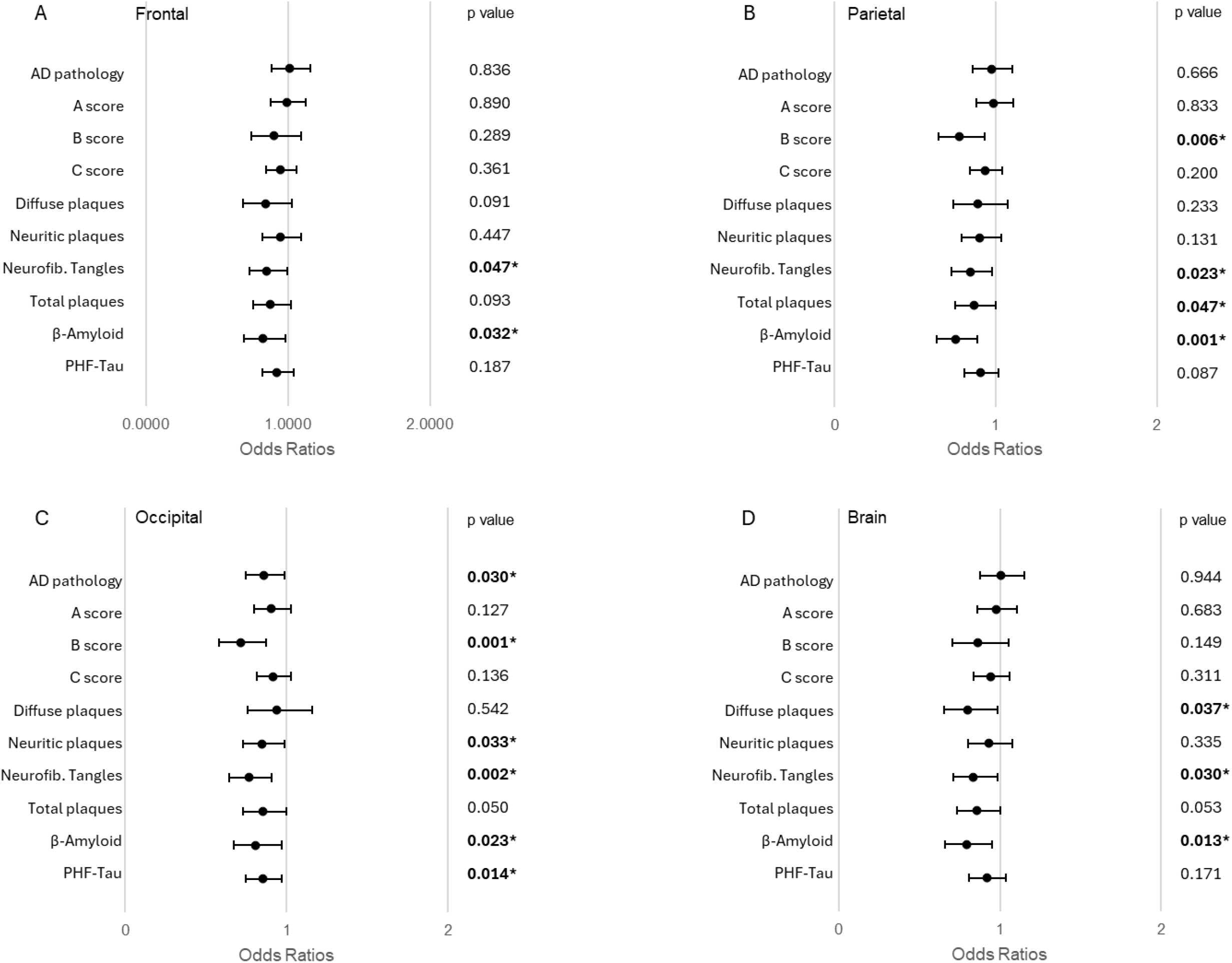
Association of AG count with neuropathologic indices. The forest plot depicts the ordinal logistic regression results (odds ratios) for AG count in different brain regions by various neuropathological indices. Findings are shown for frontal **(A)**, parietal **(B)**, occipital **(C)** and composite brain **(D)** regions. Data are representative of the entire group. All models are adjusted for age at death, sex, and years of education. Bold values indicate statistical significance (p<0.05).

### AG Size and A/T Indices

No significant associations were found between neuropathologic indices and AG size in any brain region (i.e., frontal, parietal, occipital, or whole brain; not illustrated in figures) when evaluated using regressional models, controlling for demographics.

### AG Type and A/T Indices

Next, we studied the associations of neuropathologic indices with AG type (i.e., solitary, patch, or linear) using regression models, controlling for demographics. We found that high brain β-amyloid load is associated with lower odds of patch type AG presence in the frontal region (OR, 0.80 [95% CI, 0.66–0.96]) and brain (OR, 0.76 [95% CI, 0.63–0.92]). Additionally, higher B score is associated with lower odds of patch type AG presence in the brain (OR, 0.80 [95% CI, 0.64–0.99]). There were no associations between neuropathologic indices and presence of solitary or linear AG types. Upon consideration of more quantitative variables for analysis of patch type AG, we next studied the associations of neuropathologic indices with AG patch count in different brain regions. As summarized in Fig. 3, we found that higher β-amyloid load is associated with lower odds of high AG patch count in the frontal region (OR, 0.79 [95% CI, 0.67–0.95]) and brain (OR, 0.78 [95% CI, 0.65– 0.92]); higher total plaque load is associated with lower odds of high AG patch count in the parietal region (OR, 0.83 [95% CI, 0.70–0.99]); and higher B score is associated with lower odds of high AG patch count in the brain (OR, 0.80 [95% CI, 0.66–0.96]).

**Figure 3.**
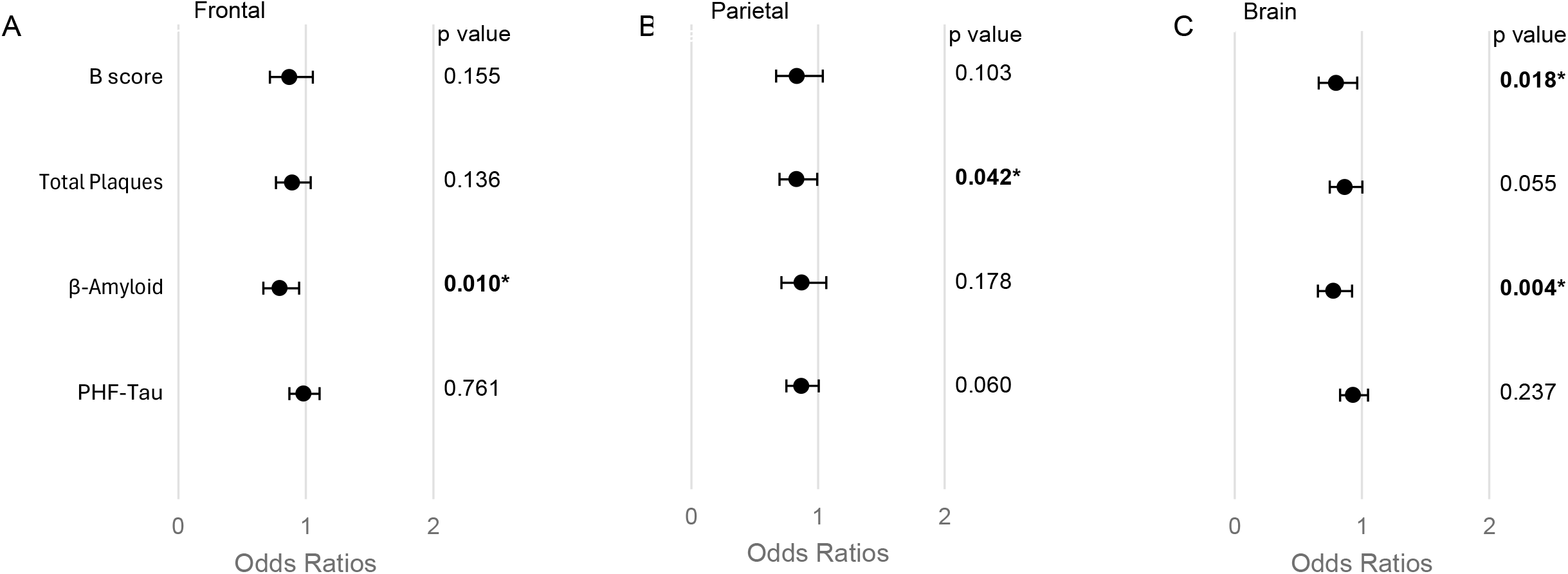
Association of AG patch count with neuropathologic indices. The forest plot depicts the ordinal logistic regression results (odds ratios) for AG patch count in different brain regions by various neuropathological indices. Findings are shown for frontal **(A)**, parietal **(B)**, and composite brain **(C)** regions. Data are representative of the entire group. All models are adjusted for age at death, sex, and years of education. Bold values indicate statistical significance (p<0.05).

### Effect of Covariates

Given the relationship of select AG properties with lobe length, brain length, and *APOE* genotype (i.e. APOE ε4 carrier status), we next studied the effects of these covariates in the main models. We found that many of the relationships between A/T indices and AG properties remained after adjusting for these covariates: B score (OR, 0.76 [95% CI, 0.61–0.94]), NFT (OR, 0.79 [95% CI, 0.66–0.94]), and Tau-IHC (OR, 0.87 [95% CI, 0.76–1.00]) remained associated with the AG number in the occipital region after adjusting for presence of *APOE* L4 allele. B score (OR, 0.68 [95% CI, 0.54–0.84]), NFT (OR, 0.74 [95% CI, 0.62–0.89]), total plaques (OR, 0.78 [95% CI, 0.66–0.93]), ADNC (OR, 0.80 [95% CI, 0.69–0.93]), β-amyloid (OR, 0.73 [95% CI, 0.60–0.90]), and PHF-Tau (OR, 0.84 [95% CI, 0.73–0.97]) remained associated with the AG number in the occipital region after adjusting for lobe length. B score (OR, 0.80 [95% CI, 0.64–1.00]), DP (OR, 0.76 [95% CI, 0.60–0.95]), total plaques (OR, 0.81 [95% CI, 0.68–0.96]), β-amyloid-IHC (OR, 0.75 [95% CI, 0.61–0.91]), PHF-Tau (OR, 0.87 [95% CI, 0.76–0.99]), and NFT (OR, 0.78 [95% CI, 0.66–0.93]) remained associated with the AG number in the brain after adjusting for brain length. B score (OR, 0.73 [95% CI, 0.58–0.92]), total plaques (OR, 0.81 [95% CI, 0.68–0.97]), β-amyloid (OR, 0.72 [95% CI, 0.58–0.89]), and PHF-Tau (OR, 0.84 [95% CI, 0.73–0.98]) remained associated with the presence of patch type AG in the brain after adjusting for brain length. B score (OR, 0.78 [95% CI, 0.62–0.93]) and β-amyloid (OR, 0.75 [95% CI, 0.63–0.91]) remained associated with the AG patch count in the brain after adjusting for lobe length.

### Interaction Effects of AG Properties on their Associations with A/T Indices

Next, we asked whether combinations of AG properties affect the associations with A/T indices. An interaction was found between frontal AG count and frontal AG size for an outcome of PHF-Tau (coef.= −0.01, *p*=0.014), whereby if decedents had higher AG count in the frontal region as well as larger AG size in the frontal region, then the increase of PHF-Tau load would be lower than the summation of the two individual effects. Additionally, an interaction was found between the brain AG patch count and brain AG size for an outcome of B score (coef.=0.03, *p*=0.043), whereby if decedents had higher AG patch count in the brain as well as larger AG size in the brain, then the increase of B score would be lower than their respective marginal associations.

Interactions were also found when stratifying by AD status (i.e., presence versus absence of intermediate to high ADNC score) [Hyman et al., 2012]. Negative interactions between frontal AG count and frontal AG size were identified, whereby if frontal AG count and frontal AG size were both larger, then the increase of PHF-Tau burden is lower than the summation of the individual effects in participants both with a pathologic diagnosis of AD (coef.= −0.02, p=0.046) and without a pathologic diagnosis of AD (coef.= −0.01, p=0.008). Additionally, there was a negative interaction between brain AG count and brain AG size, whereby if brain AG count and brain AG size were both larger, then the increase of PHF-Tau burden is lower than the summation of the individual effects in participants without a pathologic diagnosis of AD (coef.= −0.01, p=0.043). Additional interactions testing for race are summarized in Table S3. Interaction effects of race were found in the associations of AG count with various neuropathologic indices, implying that their associations were stronger in Black versus White participants.

## Discussion

We explored AG along the dorsal aspect of nearly 900 postmortem brain specimens from older adults and found that almost all brains (over 99%) exhibit presence of AG in this region. As expected, the AG were present in variable numbers, sizes, and distributions across brain specimens. AG properties were also highly heterogeneous within brains, as there was a high number of AG in the frontal brain region and gradual diminishment of the AG count along the posterior brain convexity. Overall, AG were largest in the parietal region. In regression analyses controlling for demographics, intracerebral β-amyloid load and neurofibrillary tangle counts were both inversely associated with AG count across individual and global brain regions. ADNC score, diffuse plaques, neuritic plaques, total plaques, and B score (reflecting Braak score) were also each associated with lower odds of high AG count in one or more brain regions. Despite prominent enlargement of AG with age [Shah et al., 2023], regional and global AG sizes were not found to associate with any neuropathologic A/T indices in this study.

This study also demonstrated marked heterogeneity in AG morphologic shapes. While the solitary (single or scattered) type was by far the most common type across (and within) brains, complex shapes with conglomerated patch morphology or striking arrangement into linear strands were also observed in subsets of individuals. To our knowledge the linear type, which appears as an elongated narrow stand parallel to the midsagittal plane in parasagittal dorsal brain locations, are not well described in prior literature. The origin and biological significance of the linear and patch types are unclear, and both were rare in occipital regions. Within lobar regions, the AG patch count correlated highly with the AG count, but not with AG size. Despite the absence of associations of neuropathologic indices with AG size, we found that presence of patch type AG in frontal lobe, parietal lobe, and global brain regions inversely associated with select neuropathologic indices. Specifically, in regression analyses controlling for demographics, total plaques, B score, and β-amyloid were each inversely associated with patch count in one or more brain regions. On the other hand, presence of linear type AG showed no association with any neuropathologic indices. However, it should be mentioned that due to the rarity of the linear type AG, analyses of its associations were limited by power in this study. Given available information, this study suggests that patch type AG may originate from the merging, conglomeration, or multiplication of AG. Moreover, differences in the relationships between neuropathologic indices and AG types (i.e., diffuse and patch) may suggest they have different roles in disease. Thus, further investigation of AG morphologic types is needed.

To further understand the significance of heterogeneous AG morphologies, we also tested interactions between individual AG properties and found interaction effects with respect to the outcomes of select neuropathologic A/T indices. Specifically, an interaction was found between frontal AG count and frontal AG size, whereby if decedents had higher values for both metrics, then the increase of PHF-Tau load was lower than the summation of the two individual effects. This interaction was stronger in participants without a pathologic diagnosis of AD, compared to those with AD pathologic diagnosis. Additionally, an interaction was found between brain AG patch count and brain AG size, whereby if decedents had larger values of both metrics, then the increase of B score was lower than the summation of their respective marginal associations. This interaction was also found in participants without a pathologic diagnosis of AD, but not in those with AD pathology. The biological significance of this finding requires further investigation.

We conclude that multiple AD-related neuropathologic indices inversely associate with AG count and AG patch count in one or more brain regions. Notably, intracerebral β-amyloid load (on IHC) and neurofibrillary tangle counts (on silver stain) both inversely associated with AG count in all dorsal brain regions examined, as well as in the global dorsal brain region. However, analyses suggest the need for further detailed investigation by race. Moreover, diffuse plaques and total plaques were both associated with higher odds of the more common solitary type AG. An interaction was found between frontal AG count and frontal AG size for an outcome of PHF-Tau, and this association was stronger in participants without, compared to those with a pathologic diagnosis of AD. The complexity of these associations may suggest that AG properties have a biological role in brain protein clearance, but this may vary by demographics and disease states. Thus, this study raises new questions and hypotheses. Further preclinical, clinical, and postmortem studies are needed to understand the mechanisms of the associations depicted here and to further elucidate differences in these associations across sex, race, AD status, and other factors.

This study has limitations. The participants were predominantly highly educated non-Latino White individuals above the age of 65 years [Barnes et al., 2012; Bennett et al., 2018; Schneider et al., 2009; Marquez et al., 2020]. In addition, the effects of metabolic, immune, and/or lifestyle factors were not examined. Moreover, this study did not evaluate effects of general comorbidities, which may markedly differ by race. Given the postmortem and cross-sectional nature of the analysis, cause and effect relationships cannot be drawn. Adjustments were not made for multiple comparisons since this was an exploratory study. Additionally, AG counts were measured using high-resolution postmortem digital images, therefore the study relied on 2D visualization of brain surfaces though 3D volumetric MRI analyses of AG might enhance future studies [Grossman et al., 1974]. Despite the limitations, this study also has strengths. This postmortem analysis allowed direct visualization of complex morphologic properties of AG, which may not be recognizable on MRI. The analysis also incorporated morphologic properties from almost 900 brain specimens that were harvested, processed, and evaluated in a systematic manner at a single center. Various morphologic AG properties were studied across multiple brain regions and several neuropathologic measures were incorporated in the analysis. Given the large sample size, it was possible to explore AG across race and AD status. Thus, this study should help inform the design of future hypothesis-driven studies of AG in older persons and suggests the need for consideration of demographic as well as comorbid factors.

## Abbreviations

ADNC: Alzheimer’s disease neuropathologic change
AG: arachnoid granulation
A/T: amyloid and tau
*APOE*: apolipoprotein E
PHF-Tau: paired helical filament-Tau

## Acknowledgement

The authors thank all participants in the Religious Orders Study and the Rush Memory and Aging Project. The authors also thank all Rush Alzheimer’s Disease Center staff and faculty, in particular Traci Colvin for study coordination, John Gibbons for data management, and Dominika Burba for statistical programming. This work was funded by: AARGD-22-973935, R21AG079221, P30AG10161, P30AG072975, R01AG24490, R01AG15819, R01AG017917. The funding agencies had no role in the design or conduct of these studies or the decision to submit this manuscript for publication.

