## Supplementary Material for "Arachnoid Granulation Morphologies Associate with β-Amyloid and Tau Pathology in Older Adults"

This file contains:

Extended Results

Supplementary Table 1

Supplementary Table 2

Supplementary Table 3

| | Whites (n = 746) | Blacks (n = 117) |
|---|---|---|
| **Age at death (years)** | 91.762 ±.6.100 | 84.326 ±.7.958 |
| **Education (years)** | 15.631 ± 3.138 | 14.359 ± 3.161 |
| **Brain weight (grams)** | 1171.639 ± 576.309 | 1084.296 ± 125.986 |
| **Postmortem interval (hours)** | 9.436 ± 8.472 | 16.799 ± 17.100 |
| ***APOE* Ɛ4 allele:** | 160 (21.45%) | 46 (39.32%) |
| **Lobe Length (mm):** Frontal | 145.664 ± 10.638 | 142.943 ± 11.201 |
| Parietal | 76.247 ± 8.735 | 74.818 ± 10.941 |
| Occipital | 54.975 ± 5.954 | 53.114 ± 6.884 |
| Cerebellum | 64.011 ± 5.515 | 61.337 ± 5.187 |
| **Pathologic Diagnosis of AD:** | | |
| None to Mild ADNC | 268 (35.92%) | 51 (43.59%) |
| Moderate to Severe ADNC | 478 (64.08%) | 66 (56.41%) |
| **ADNC Score:** | | |
| A Score | 1.928 ± 1.052 | 1.696 ± 1.086 |
| B Score | 2.142 ± 0.658 | 1.957 ± 0.736 |
| C Score | 1.732 ± 1.122 | 1.624 ± 1.230 |
| **ADNC Lesion:** | | |
| Diffuse Plaques | 0.647 ± 0.630 | 0.677 ± 0.746 |
| Neuritic Plaques | 0.850 ± 0.842 | 0.842 ± 0.842 |
| Neurofibrillary Tangles | 0.705 ± 0.821 | 0.658 ± 0.836 |
| Total Plaques | 0.999 ± 0.853 | 0.940 ± 0.910 |
| B-Amyloid (IHC) | 1.016 ± 0.726 | 0.887 ± 0.712 |
| PHF-Tau (IHC) | 1.408 ± 1.032 | 1.236 ± 1.135 |

**Supplemental Table 1.** Demographic and neuropathologic characteristics of participants and postmortem brain specimens by race (n = 882 in total).

| | **Whites** (n = 746) | **Blacks** (n = 117) |
|---|---|---|
| **Brains with any AG (n, %)** | | |
| Frontal | 721 (96.65%) | 111 (94.87%) |
| Parietal | 712 (95.44%) | 109 (93.16%) |
| Occipital | 656 (87.94%) | 97 (82.91%) |
| Brain* | 745 (99.87 %) | 116 (99.15%) |
| **AG count**** | | |
| Frontal | 3.960 ± 1.373 | 3.675 ± 1.436 |
| Parietal | 3.160 ± 1.331 | 3.085 ± 1.492 |
| Occipital | 0.932 ± 0.610 | 0.944 ± 0.771 |
| Brain* | 2.082 ± 0.662 | 1.998 ± 0.714 |
| **AG size (max, in mm)** | | |
| Frontal | 15.513 ± 8.238 | 13.912 ± 7.523 |
| Parietal | 12.264 ± 6.860 | 10.589 ± 6.639 |
| Occipital | 9.255 ± 6.276 | 5.636 ± 3.355 |
| Brain* | 16.980 ± 8.104 | 15.069 ± 7.604 |
| **Presence of diffuse type AG (n, %)** | | |
| Frontal | 719 (96.38%) | 112 (95.73%) |
| Parietal | 710 (95.17%) | 109 (93.16%) |
| Occipital | 654 (87.67%) | 99 (84.62%) |
| **Presence of linear type AG (n, %)** | | |
| Frontal | 148 (19.84%) | 18 (15.38%) |
| Parietal | 81 (10.86%) | 16 (13.68%) |
| Occipital | 2 (0.27%) | 1 (0.85%) |
| **Presence of patch type AG (n, %)** | | |
| Frontal | 426 (57.10%) | 52 (44.44%) |
| Parietal | 222 (29.76%) | 38 (32.48%) |
| Occipital | 28 (3.75%) | 3 (2.56%) |
| **AG Patch Count (n)** | | |
| Frontal | 0.81 ± 0.77 | 0.64 ± 0.76 |
| Parietal | 0.39 ± 0.62 | 0.39 ± 0.56 |
| Occipital | 0.04 ± 0.20 | 0.04 ± 0.19 |
| Brain | 0.90 ± 0.77 | 0.76 ± 0.73 |

**Supplemental Table 2.** AG characteristics in postmortem brain specimens, summarized by race (n = 882 in total). *"Brain" represents composite dorsal cerebrum. **AG counts represent the mean of semiquantitative scores from the left and right sides.

| Interactions | Tested In the Entire Group | | | | | | | | |
|---|---|---|---|---|---|---|---|---|---|
| Outcome: | ADNC Score | A Score | C Score | Diffuse Plaques | Neuritic Plaques | Total Plaques | NFT/ Neuritic Plaques | β-Amyloid (IHC) | PHF-Tau (IHC) |
| **Model 1:** diffuse (F)* race | **2.29<br>0.046*** | **2.44<br>0.034*** | 2.17<br>0.066 | 0.49<br>0.138 | 0.72<br>0.110 | 0.70<br>0.109 | 0.36<br>0.356 | 0.49<br>0.177 | -0.26<br>0.622 |
| **Model 2:** linear (F)* race | 0.71<br>0.154 | **1.04<br>0.045*** | 0.75<br>0.137 | **0.39<br>0.036*** | 0.34<br>0.154 | 0.47<br>0.056 | 0.29<br>0.168 | 0.22<br>0.268 | 0.15<br>0.599 |
| **Model 3:** patch (F)* race | 0.50<br>0.175 | 0.59<br>0.109 | **0.94<br>0.011*** | 0.10<br>0.458 | **0.39<br>0.030*** | 0.19<br>0.287 | **0.35<br>0.025*** | 0.25<br>0.090 | 1.27<br>0.193 |
| Model 4: diffuse (P)* race | 0.68<br>0.363 | 1.11<br>0.138 | 1.43<br>0.073 | 0.30<br>0.261 | 0.61<br>0.092 | 0.54<br>0.127 | 0.52<br>0.099 | 0.42<br>0.156 | 0.55<br>0.192 |
| **Model 5:** linear (P)* race | **1.21<br>0.025*** | **1.49<br>0.007*** | 0.91<br>0.091 | **0.59<br>0.003*** | 0.48<br>0.063 | **0.75<br>0.004*** | 0.26<br>0.247 | 0.34<br>0.119 | -0.01<br>0.968 |
| Model 6: patch (P)* race | -0.03<br>0.939 | -0.05<br>0.902 | 0.17<br>0.669 | -0.11<br>0.437 | 0.09<br>0.634 | -0.01<br>0.944 | 0.10<br>0.528 | 0.03<br>0.830 | 0.24<br>0.294 |
| **Model 8:** linear (B)* race | 0.87<br>0.041 | **1.09<br>0.012*** | 0.71<br>0.097 | **0.36<br>0.023*** | 0.35<br>0.093 | **0.41<br>0.047*** | 0.22<br>0.222 | 0.28<br>0.104 | 0.12<br>0.620 |
| **Model 9:** patch (B)* race | 0.09<br>0.816 | 0.21<br>0.569 | 0.33<br>0.371 | 0.05<br>0.686 | 0.17<br>0.354 | 0.10<br>0.559 | 0.13<br>0.399 | 0.11<br>0.453 | -0.01<br>0.949 |
| **Model 10:** AG count (F)* race | 0.**50<br>0.009*** | **0.61<br>0.001*** | **0.52<br>0.006*** | **0.14<br>0.038*** | **0.28<br>0.002*** | **0.25<br>0.006*** | **0.22<br>0.007*** | **0.19<br>0.009*** | 0.12<br>0.264 |
| **Model 11:** AG count (P)* race | **0.40<br>0.005*** | **0.49<br><0.001*** | **0.46<br>0.002*** | **0.13<br>0.009*** | **0.21<br>0.001*** | **0.19<br>0.004*** | **0.16<br>0.007*** | **0.15<br>0.008*** | 0.03<br>0.751 |
| **Model 12:** AG count (O)* race | 0.16<br>0.215 | 0.23<br>0.077 | **0.28<br>0.032*** | 0.06<br>0.178 | **0.15<br>0.020*** | **0.13<br>0.029*** | 0.10<br>0.057 | 0.09<br>0.079 | 0.06<br>0.401 |
| **Model 13:** AG count (B)* race | 0.39<br>0.059 | **0.44<br>0.036*** | **0.49<br>0.021*** | **0.17<br>0.032*** | **0.23<br>0.021*** | 0.18<br>0.087 | **0.18<br>0.041*** | **0.18<br>0.024*** | **0.30<br>0.012*** |

**Supplemental Table 3. Interaction effects of AG properties and race on their associations with various neurodegenerative indices**. Results are shown for various neurodegenerative scores and lesions (outcomes in row one). As shown, select AG properties interact with race and suggest that effects of AG size, count, and patch count (in a given brain region) on neurodegenerative indices may differ depending on race. Values represent coefficients (top line) or p values (bottom line). Bold values indicate statistical significance (p<0.05). Abbreviations: B, brain; F, frontal; N, neurofibrillary tangles; P, parietal; PHF, paired helical filament; O, occipital.
